# FDA-authorized COVID-19 vaccines are effective per real-world evidence synthesized across a multi-state health system

**DOI:** 10.1101/2021.02.15.21251623

**Authors:** Colin Pawlowski, Patrick Lenehan, Arjun Puranik, Vineet Agarwal, AJ Venkatakrishnan, Michiel J.M. Niesen, John C. O’Horo, Abinash Virk, Melanie D. Swift, Andrew D. Badley, John Halamka, Venky Soundararajan

**Author notes:** Address correspondence to Venky Soundararajan. Joint first authors (Colin Pawlowski, Patrick Lenehan).

## Abstract

Large Phase 3 clinical trials of the two FDA-authorized COVID-19 vaccines, mRNA-1273 (Moderna) and BNT162b2 (Pfizer/BioNTech), have demonstrated efficacies of 94.1% (n = 30,420, 95% CI: 89.3-96.8) and 95% (n = 43,448, 95% CI: 90.3-97.6) in preventing symptomatic COVID-19, respectively. Given the ongoing vaccine rollout to healthcare personnel and residents of long-term care facilities, here we provide a preliminary assessment of real-world vaccination efficacy in 62,138 individuals from the Mayo Clinic and associated health system (Arizona, Florida, Minnesota, Wisconsin) between December 1^st^ 2020 and February 8^th^ 2021. Our retrospective analysis contrasts 31,069 individuals receiving at least one dose of either vaccine with 31,069 unvaccinated individuals who are propensity-matched based on demographics, location (zip code), and number of prior SARS-CoV-2 PCR tests. 8,041 individuals received two doses of a COVID-19 vaccine and were at risk for infection at least 36 days after their first dose. Administration of two COVID-19 vaccine doses was 88.7% effective in preventing SARS-CoV-2 infection (95% CI: 68.4-97.1%) with onset at least 36 days after the first dose. Furthermore, vaccinated patients who were subsequently diagnosed with COVID-19 had significantly lower 14-day hospital admission rates than propensity-matched unvaccinated COVID-19 patients (3.7% vs. 9.2%; Relative Risk: 0.4; p-value: 0.007). Building upon the previous randomized trials of these vaccines, this study demonstrates their real-world effectiveness in reducing the rates of SARS-CoV-2 infection and COVID-19 severity among individuals at highest risk for infection.

## Introduction

To date there have been over 107 million confirmed cases of COVID-19 and over 2.3 million associated deaths globally (1). From the moment that SARS-CoV-2 was identified as the causative agent of COVID-19, efforts were initiated to characterize this virus and to develop vaccines against it (2, 3, 4). Within months, several candidates were shown to be safe and to induce robust immune responses against SARS-CoV-2 in a series of early phase trials (5, 6, 7, 8). More recently, multiple vaccine candidates have shown over 94% efficacy in preventing symptomatic COVID-19 infection in large phase 3 clinical trials (9, 10). Of note, unlike the seasonal flu vaccine, both of these candidates are delivered as a series of two inoculations separated by three or four weeks, with maximal response believed to be achieved by one to two weeks after the second dose (5, 6, 9, 10).

In a phase 3 trial studying BNT162b2 (9), the COVID-19 vaccine candidate developed by Pfizer/BioNTech, 50 out of 21,314 (0.23%) vaccinated patients experienced a symptomatic COVID-19 infection, with an incidence rate of 12.5 cases per 1000 person-years. In contrast, 275 of 21,258 (1.29%) patients receiving a placebo injection developed COVID-19, with an incidence rate of 69.1 cases per 1000 person-years. Thirty patients experienced severe disease, all of whom had received placebo. Seven or more days after the second dose, the difference between groups was even more pronounced, with incidence rates of 3.61 and 72.9 cases per 1000 person-years in the vaccinated and placebo groups, respectively (efficacy = 95.0%; 95% CI: 90.3-97.6%).

Similarly, in the trial studying mRNA-1273 (10), the vaccine candidate developed by Moderna, 19 of 14,550 (0.13%) vaccinated patients experienced a symptomatic infection compared to 269 of 14,598 (1.84%) patients receiving placebo. Among these symptomatic infections, there were 9 cases of severe COVID-19 in the placebo group compared to only one in the vaccinated cohort. This effect was stronger when considering infection rates 14 or more days after the second dose, with incidence rates of 3.3 and 56.5 cases per 1000 person-years in the vaccinated and placebo groups, respectively (efficacy = 94.1%; 95% CI: 89.3-96.8%).

Both BNT162b2 and mRNA-1273 are now being administered throughout the United States under an Emergency Use Authorization by the FDA, with first priority given to individuals at high risk for becoming infected with SARS-CoV-2 or experiencing severe COVID-19, including healthcare workers and residents of long term care facilities (11). While these groups were not excluded from the phase 3 trials, vaccine efficacy has not been specifically demonstrated among them. It is thus critical to analyze outcomes of vaccinated patients to date to determine whether these vaccines are indeed effective in especially high-risk individuals.

Here we conducted a large-scale real world interim analysis of COVID-19 vaccination outcomes in the United States. Specifically, we assessed the rates of SARS-CoV-2 positivity and severity of COVID-19 among 31,069 individuals who received at least one dose of BNT162b2 or mRNA-1273 in the Mayo Clinic health system, including sites in Minnesota, Arizona, Florida, and Wisconsin. One challenge inherent to such real world analyses is the lack of a built-in placebo arm, which is essential to establish the expected infection rate during the study period and thereby to assess vaccine efficacy. To address this shortcoming, we used 1-to-1 propensity score matching to generate a cohort of 31,069 individuals who were not previously infected with SARS-CoV-2 and did not receive a COVID-19 vaccine by the end of the study period (**Figure 1, Table 1**). This control arm was balanced for potential confounding factors for COVID-19 infection, including: demographics, location (zip code), and number of PCR tests received prior to the study period. Using this control arm, we compared rates of SARS-CoV-2 infection during defined intervals after study enrollment.

**Table 1.** Clinical characteristics of vaccinated and 1:1 propensity-matched unvaccinated cohorts. Covariates for balancing include: (1) Demographics (age, sex, race, ethnicity), (2) Number of prior PCR tests (number of PCR tests that the individual received before December 1, 2020), and (3) Location (zip code). Note that the zip code is matched exactly between the two cohorts, so the proportion of individuals in each state is identical. Highly balanced covariates with Standardized Mean Difference (SMD) < 0.1 are indicated with ***.

| Clinical covariate | Vaccinated cohort | 1:1 Propensity-matched unvaccinated cohort | Standardized Mean Difference (SMD) |
| --- | --- | --- | --- |
| Total number of patients | 31,069 | 31,069 |  |
| Age in years |  |  |  |
| - 18-24 | 1,288 (4.1%) | 1,446 (4.7%) | 0.02*** |
| - 25-34 | 5,668 (18.2%) | 5,382 (17.3%) | 0.02*** |
| - 35-44 | 5,344 (17.2%) | 5,311 (17.1%) | 0.00*** |
| - 45-54 | 3,980 (12.8%) | 4,131 (13.3%) | 0.01*** |
| - 55-64 | 4,288 (13.8%) | 4,426 (14.2%) | 0.01*** |
| - 65-74 | 3,398 (10.9%) | 4,010 (12.9%) | 0.06*** |
| - 75+ | 7,103 (22.9%) | 6,363 (20.5%) | 0.06*** |
| Sex |  |  |  |
| - Female | 19,321 (62.2%) | 20,105 (64.7%) | 0.05*** |
| - Male | 11,743 (37.8%) | 10,958 (35.3%) | 0.05*** |
| - Unknown | 5 (0.0%) | 6 (0.0%) | 0.00*** |
| Race |  |  |  |
| - Asian | 1,306 (4.2%) | 1,213 (3.9%) | 0.02*** |
| - Black / African American | 649 (2.1%) | 610 (2.0%) | 0.01*** |
| - Native American | 75 (0.2%) | 60 (0.2%) | 0.01*** |
| - White / Caucasian | 27,987 (90.1%) | 28,140 (90.6%) | 0.02*** |
| - Other | 660 (2.1%) | 654 (2.1%) | 0.00*** |
| - Unknown | 392 (1.3%) | 392 (1.3%) | 0.00*** |
| Ethnicity |  |  |  |
| - Hispanic or Latino | 949 (3.1%) | 987 (3.2%) | 0.01*** |
| - Not Hispanic or Latino | 29,257 (94.2%) | 29,214 (94.0%) | 0.01*** |
| - Unknown | 863 (2.8%) | 868 (2.8%) | 0.00*** |
| Number of prior PCR tests |  |  |  |
| - 0 | 4,542 (14.6%) | 4,467 (14.4%) | 0.01*** |
| - 1 | 13,280 (42.7%) | 14,554 (46.8%) | 0.08*** |
| - 2 | 6,601 (21.2%) | 6,247 (20.1%) | 0.03*** |
| - 3 | 3,296 (10.6%) | 2,735 (8.8%) | 0.06*** |
| - 4 | 1,619 (5.2%) | 1,338 (4.3%) | 0.04*** |
| - 5+ | 1,731 (5.6%) | 1,728 (5.6%) | 0.00*** |
| State |  |  |  |
| - Arizona | 1,627 (5.2%) | 1,627 (5.2%) | 0.00*** |
| - Florida | 5,581 (18.0%) | 5,581 (18.0%) | 0.00*** |
| - Minnesota | 18,236 (58.7%) | 18,236 (58.7%) | 0.00*** |
| - Wisconsin | 5,625 (18.1%) | 5,625 (18.1%) | 0.00*** |

**Figure 1.**
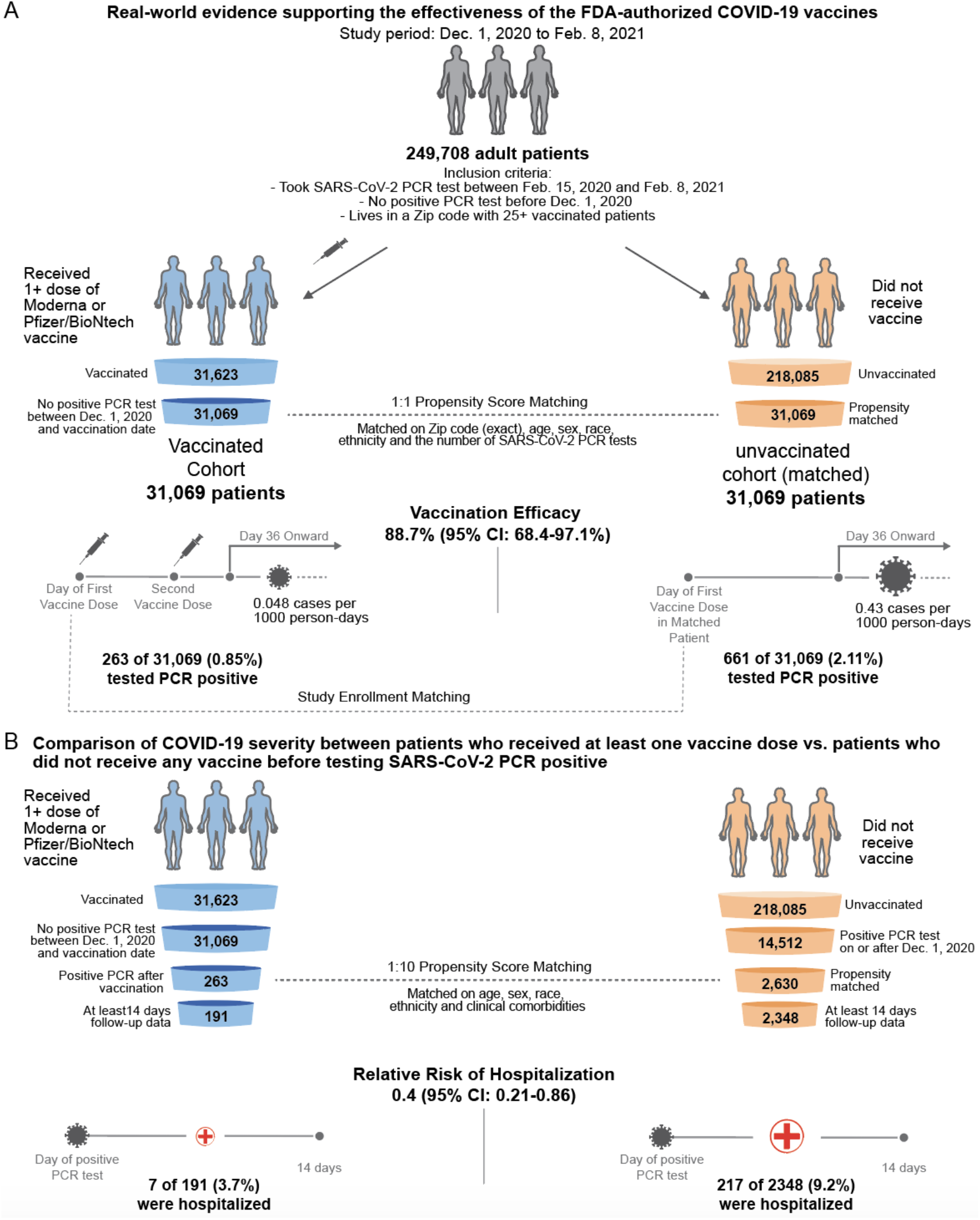
Schematic illustrating the algorithms for participant selection and outcome assessment. (A) Design of study to compare SARS-CoV-2 infection rates in patients receiving COVID-19 vaccination compared to 1-to-1 propensity matched unvaccinated patients (n = 31,069 per group). For each group, incidence rates were calculated to assess the efficacy of vaccination in preventing SARS-CoV-2 infection, as defined by a positive PCR test, with onset at least 36 days after the first dose or the date of study enrollment. Several other time windows were also evaluated for vaccine efficacy. (B) Design of study to compare COVID-19 disease severity in patients who were vaccinated prior to diagnosis with COVID-19 and had at least 14 days of follow-up after diagnosis (n = 191) versus 1-to-10 propensity matched unvaccinated patients with at least 14 days of follow-up (n = 2,348). Severity outcomes (hospitalization, ICU admission, and mortality) were assessed within 14 days of PCR diagnosis.

In addition, in order to evaluate the potential impact of COVID-19 vaccination upon disease severity, we conducted a similar matched analysis on the 263 vaccinated individuals who subsequently tested positive for SARS-CoV-2 during the study period following their first dose of vaccine. In particular, we used 1-to-10 propensity score matching to generate a cohort of 2,630 individuals who tested positive for SARS-CoV-2 during the study period who never received any vaccination for COVID-19. This control arm was balanced for potential confounding factors for severity of COVID-19 illness, including demographics and comorbidities identified from the clinical notes in the Mayo Clinic electronic health records for these patients. Using this control arm, we compared rates of hospital admission, ICU admission, and mortality in the 14 days following the date of the SARS-CoV-2 infection.

## Methods

### Study design, setting and population

This is a retrospective study of individuals who underwent polymerase chain reaction (PCR) testing for suspected SARS-CoV-2 infection at the Mayo Clinic and hospitals affiliated to the Mayo health system. This study was reviewed and approved by the Mayo Clinic Institutional Review Board (IRB 20-003278) as a minimal risk study. Subjects were excluded if they did not have a research authorization on file. The IRB approved was titled: Study of COVID-19 patient characteristics with augmented curation of Electronic Health Records (EHR) to inform strategic and operational decisions with the Mayo Clinic.

In total, there were 507,525 patients in the Mayo electronic health record (EHR) database who received a PCR test between February 15, 2020 and February 8, 2021. To obtain the study population, we defined the following inclusion criteria: (1) at least 18 years old; (2) no positive SARS-CoV-2 PCR test before December 1, 2020; (3) resides in a locale (based on Zip code) with at least 25 patients who have received BNT162b2 or mRNA-1273. This population included 249,708 patients, of whom 31,623 have received BNT162b2 or mRNA-1273 and 218,085 have no record of COVID-19 vaccination. Vaccinated individuals who had tested positive for SARS-CoV-2 by PCR between December 1, 2020 and the date of their first vaccine dose were excluded, resulting in 31,299 individuals. Individuals with zero follow-up days after vaccination (i.e. those who received the first vaccine dose on the date of data collection) were also excluded, leaving 31,069 patients in the final vaccinated cohort. Propensity matched unvaccinated cohorts for analyses of vaccine efficacy were selected from the previously derived set of 218,085 unvaccinated patients. This patient selection algorithm and its associated counts are summarized in **Figure 1A**. More details on the propensity score matching procedure for the vaccine efficacy analysis are provided in the next section.

We conducted a similar matching analysis to assess the impact of vaccination upon COVID-19 disease severity. For this analysis, we considered the 263 vaccinated patients who tested positive for SARS-CoV-2 following their first vaccine dose during the study period, and 14,512 unvaccinated patients who tested positive for SARS-CoV-2 during the study period. For each of the 263 SARS-CoV-2 positive vaccinated patients, we selected 10 controls from the unvaccinated cohort using 1-to-10 propensity score matching. This patient selection algorithm and its associated counts are summarized in **Figure 1B**. More details on the propensity score matching procedures for the disease severity analysis are provided in a subsequent section.

### Propensity score matching to select the unvaccinated cohort for efficacy analysis

We employed 1:1 propensity score matching (12) to construct an unvaccinated cohort similar to the vaccinated cohort on key risk factors for SARS-CoV-2 infection, including geography, demographics, and records of PCR testing. Specifically, first we matched exactly based on geography (zip code of the patient’s residence). Next, we used propensity score matching to match approximately based upon demographic features (age, sex, race, ethnicity) and records of PCR testing (number of negative PCR tests taken between February 8, 2020 and November 30, 2020); the number of negative PCR tests covariate serves as a proxy for ongoing baseline exposure to COVID. To obtain the propensity scores for the matching procedure, we trained regularized logistic regression models for each zip code using the software package sklearn v0.20.3 in Python.

Using these propensity scores, we then matched each of the 31,299 vaccinated patients (who received at least one dose of BNT162b2 or mRNA-1273 and had not tested positive for SARS-CoV-2 between December 1, 2020 and the date of their first vaccination) with 1 patient out of the 218,085 unvaccinated patients, using greedy nearest-neighbor matching without replacement (13). In particular, for each vaccinated patient, we selected an unvaccinated patient that lived in the same zip code with the closest propensity score to the vaccinated patient. The cohorts were then subsetted to only include the 31,069 vaccinated individuals (and their corresponding propensity matched individuals) who had at least one day of follow-up after their date of study enrollment.

For each vaccinated patient, the date of study enrollment (Day 0) was defined as the date of their first vaccine dose. For each unvaccinated patient, the date of study enrollment was defined as the date of the first vaccine dose for their matched vaccinated patient. The resulting cohorts are summarized in **Table 1** along with the standardized mean differences (SMD) of their clinical covariates (14, 15). Overall, there is no substantial difference between the two cohorts in any of the clinical covariates that were included in propensity score matching (with SMD < 0.1 for all covariates). The age distributions of patients in the vaccinated, unmatched unvaccinated, and propensity matched unvaccinated cohorts are shown in **Figure S1A-B**. Additional data regarding the mean follow-up time, the number of vaccine doses received, and the number of patients taking at least one SARS-CoV-2 PCR test after the study enrollment date are provided in **Table S1**. The distribution of total follow-up time (i.e. days between first vaccine dose and the end of the data collection period) for the 31,299 vaccinated patients is provided in **Figure S2A**.

### Evaluation of vaccine efficacy

To evaluate the COVID-19 vaccine efficacy in a real-world clinical setting, we compared the two populations described above and summarized in **Figure 1**: (1) 31,069 individuals with follow-up who received BNT162b2 or mRNA-1273 and did not have a prior positive SARS-CoV-2 PCR test (“vaccinated”), and (2) 31,069 propensity matched individuals who have never received a COVID-19 vaccine and did not have a positive SARS-CoV-2 PCR test before the first vaccination date (dose 1) of their matched patient (“unvaccinated”).

Cumulative proportional incidence of SARS-CoV-2 infection was compared between vaccinated and unvaccinated patients by Kaplan Meier analysis. Cumulative proportional incidence at time *t* is the estimated proportion of patients who experience the outcome on or before time *t*, i.e. 1 minus the standard Kaplan-Meier survival estimate. We considered cumulative incidence starting at Day 1, Day 14, and Day 28 relative to the date of study enrollment (Day 0). Statistical significance was assessed with the log rank test (16).

Efficacy was also assessed during defined intervals by computing the incidence rate ratio (IRR) of the vaccinated and unvaccinated cohorts. Efficacy was defined as 100% x (1 -IRR). The time periods considered were as follows: (1) Day 1 onwards, (2) Day 15 onwards, (3) Day 29 onwards, (4) Day 36 onwards, and (5) six one-week intervals starting one day after the first dose of vaccination (“Day 1”). Only six one-week intervals were considered because less than 25% of the patients in each cohort had adequate follow-up to contribute at-risk person days after this time. For each cohort in a given time period, incidence rates were calculated as the number of patients testing positive for SARS-CoV-2 in that time period divided by the total number of at-risk person-days contributed in that time period. For each patient, at-risk person-days are defined as the number of days in the time period in which the patient has not yet tested positive for SARS-CoV-2 or died. The IRR was calculated as the incidence rate of the vaccinated cohort divided by the incidence rate of the unvaccinated cohort, and its 95% confidence interval was computed using an exact approach described previously (17).

### Propensity score matching to select the unvaccinated COVID-19 patients for disease severity analysis

Similarly, we applied 1:10 propensity score matching (12) to construct a SARS-CoV-2 positive unvaccinated cohort similar in baseline clinical covariates to the cohort of patients who were vaccinated and subsequently tested positive for SARS-CoV-2. In particular, we used propensity score matching to match approximately based upon demographic features (age, sex, race, ethnicity) and comorbidities (asthma, cancer, cardiomyopathy, chronic kidney disease, chronic obstructive pulmonary disease, coronary artery disease, heart failure, hypertension, obesity, pregnancy, severe obesity, sickle cell disease, solid organ transplant, stroke / cerebrovascular disease, type 2 diabetes mellitus). This list of comorbidities was derived from the list of risk factors for severe COVID-19 illness provided by the Centers of Disease Control and Prevention (18). We used deep neural networks to automatically identify comorbidities from the clinical notes, which are described in the next section. To obtain the propensity scores, we trained a regularized logistic regression model with these features using the software package sklearn v0.20.3 in Python.

Based on these propensity scores, we matched each of the 263 individuals that tested positive for SARS-CoV-2 after vaccination with 10 individuals out of the 14,512 individuals that tested positive for SARS-CoV-2 and that were not vaccinated. As in the previous propensity score matching procedure, we used greedy nearest-neighbor matching without replacement (13). The resulting cohorts are summarized in **Table 3**, along with the SMDs for the clinical covariates that were balanced upon (14, 15). Overall, there is no significant difference between the two cohorts in any of the clinical covariates that were included in propensity score matching (with SMD < 0.1 for all covariates). The age distributions of COVID-19 patients in the vaccinated cohort, unmatched unvaccinated cohort, and propensity matched unvaccinated cohort are shown in **Figure S1C-D**. The distribution of total follow-up time (i.e. days between diagnosis date and the end of the data collection period) is provided in **Figure S2B**.

**Table 3.** Clinical characteristics of SARS-CoV-2 positive vaccinated and 1:10 propensity matched unvaccinated cohorts. The SARS-CoV-2 positive vaccinated cohort includes all patients who received at least one dose of a COVID-19 vaccine and then subsequently received a positive PCR test. The control cohort is a 1:10 propensity-matched cohort derived from the set of unvaccinated patients with a positive PCR test on or after December 1, 2020. Demographics and comorbidities are presented for each cohort, and number of doses is presented for the vaccinated cohort. Comorbidities were determined via neural network models applied to clinical notes for each patient between December 1, 2015 and November 30, 2020. Highly balanced covariates with Standardized Mean Difference (SMD) < 0.1 are indicated with ***.

| Clinical covariate | SARS-CoV-2 positive vaccinated cohort | 1:10 Propensity-matched SARS-CoV-2 positive unvaccinated cohort | Standardized Mean Difference (SMD) |
| --- | --- | --- | --- |
| Total number of patients | 263 | 2630 |  |
| Age |  |  |  |
| - 18-44 years old | 110 (41.8%) | 1055 (40.1%) | 0.03*** |
| - 45 - 64 years old | 103 (39.2%) | 1008 (38.3%) | 0.02*** |
| - >= 65 years old | 50 (19.0%) | 567 (21.6%) | 0.06*** |
| Sex |  |  |  |
| - Female | 179 (68.1%) | 1786 (67.9%) | 0.00*** |
| - Male | 84 (31.9%) | 844 (32.1%) | 0.00*** |
| - Unknown | 0 (0.0%) | 0 (0.0%) | N/A |
| Race |  |  |  |
| - Asian | 12 (4.6%) | 121 (4.6%) | 0.07*** |
| - Black / African American | 2 (0.8%) | 16 (0.6%) | 0.01*** |
| - Native American | 0 (0%) | 1 (0.04%) | 0.02*** |
| - White / Caucasian | 243 (92.4%) | 2454 (93.3%) | 0.09*** |
| - Other | 4 (1.5%) | 31 (1.2%) | 0.06*** |
| - Unknown | 2 (0.8%) | 7 (0.3%) | 0.07*** |
| Ethnicity |  |  |  |
| - Hispanic or Latino | 9 (3.4%) | 77 (2.9%) | 0.03*** |
| - Not Hispanic or Latino | 247 (93.9%) | 2492 (94.8%) | 0.04*** |
| - Unknown | 7 (2.7%) | 61 (2.3%) | 0.02*** |
| Comorbidities |  |  |  |
| - Asthma | 35 (13.3%) | 388 (14.8%) | 0.04*** |
| - Cancer | 64 (24.3%) | 708 (26.9%) | 0.06*** |
| - Cardiomyopathy | 5 (1.9%) | 49 (1.9%) | 0.00*** |
| - Chronic Kidney Disease | 15 (5.7%) | 157 (6.0%) | 0.01*** |
| - Chronic Obstructive Pulmonary Disease | 11 (4.2%) | 119 (4.5%) | 0.02*** |
| - Coronary Artery Disease | 19 (7.2%) | 201 (7.6%) | 0.02*** |
| - Heart failure | 12 (4.6%) | 134 (5.1%) | 0.02*** |
| - Hypertension | 69 (26.2%) | 751 (28.6%) | 0.05*** |
| - Obesity | 57 (21.7%) | 572 (21.7%) | 0.00*** |
| - Pregnancy | 1 (0.4%) | 7 (0.3%) | 0.02*** |
| - Severe Obesity | 8 (3.0%) | 70 (2.7%) | 0.02*** |
| - Sickle Cell Disease | 0 (0.0%) | 0 (0.0%) | N/A |
| - Solid Organ Transplant | 0 (0.0%) | 0 (0.0%) | N/A |
| - Stroke / Cerebrovascular Disease | 1 (0.4%) | 7 (0.3%) | 0.02*** |
| - Type 2 Diabetes Mellitus | 29 (11.0%) | 292 (11.1%) | 0.00*** |
| Number of vaccine doses |  |  |  |
| - 1 dose | 164 (62.3%) |  |  |
| - 2 doses | 99 (37.6%) |  |  |

For each SARS-CoV-2 positive patient in both the vaccinated and unvaccinated cohorts, the index date for the analysis (day 0) was taken to be the date of the first positive PCR test. Clinical outcomes at 14 days were compared, including hospital admission, ICU admission, and mortality.

### Evaluation of disease severity

We compare the clinical outcomes of vaccinated and propensity-matched unvaccinated SARS-CoV-2 positive patients in order to evaluate the impact of COVID-19 vaccination upon disease severity. Among the patients in each cohort with at least 14 days of follow-up after their first positive PCR test (n = 191 vaccinated, 2,348 unvaccinated), we evaluate: (1) 14-day hospital admission rate: Number of patients admitted to the hospital in the two weeks following their positive PCR test, (2) 14-day ICU admission rate: Number of patients admitted to the ICU in the two weeks following their positive PCR test, and (3) 14-day mortality rate: Number of patients deceased in the two weeks following their positive PCR test. For each outcome, we report the relative risk (rate in the vaccinated cohort divided by the rate in the matched unvaccinated cohort), 95% confidence interval for the relative risk (19), and Fisher’s exact test p-value. Hospital-free and ICU-free survival were also compared via Kaplan-Meier analysis, with statistical significance assessed with the log rank test (16).

### Deep neural networks to identify comorbidities from clinical notes

In order to identify the comorbidities from the electronic health record for each patient, we used a BERT-based neural network model (20) to classify the sentiment for the phenotypes that appeared in the clinical notes. In particular, we applied a phenotype sentiment classification model that had been trained on 18,500 sentences which achieves an out-of-sample accuracy of 93.6% with precision and recall scores above 95% (21). This classification model predicts four classes, including: (1) “Yes”: confirmed diagnosis (2) “No”: ruled-out diagnosis, (3) “Maybe”: possibility of disease, and (4) “Other”: alternate context (e.g. family history of disease). For each patient, we applied the sentiment model to the clinical notes in the Mayo Clinic electronic health record from December 1, 2015 to November 30, 2020. For each comorbidity phenotype, if a patient had at least one mention of the phenotype during the time period with a confidence score of 90% or greater, then the patient was labelled as having the phenotype.

## Results

### COVID-19 vaccines reduce the incidence rate of SARS-CoV-2 infection

Over the duration of our study (see **Methods**), 263 of 31,069 (0.85%) vaccinated individuals tested positive for SARS-CoV-2 compared to 661 of 31,069 (2.13%) matched unvaccinated individuals (**Table 2, Figure 1**). The incidence rates of positive SARS-CoV-2 tests in the vaccinated and unvaccinated cohorts were 0.31 and 0.8 cases per 1000 person-days, respectively. This corresponds to a vaccine efficacy of 60.7% (95% CI: 54.6-66.1%) over the entire study period, and a log-rank test indicates that the hazard rate is significantly lower in the vaccinated cohort over this time interval (p = 5×10^−40^; **Figure 2A**). The hazard rates were also significantly lower in the vaccinated group when considering SARS-CoV-2 infections with onset at 14 days after study enrollment (p = 1×10^−28^; **Figure 2B**) or 28 days after study enrollment (p = 2.4×10^−13^; **Figure 2C**). For the 263 vaccinated individuals who subsequently tested positive for SARS-CoV-2, the distribution of time from first dose to first positive SARS-CoV-2 PCR test is shown in **Figure 3**.

**Table 2.** SARS-CoV-2 incidence rates in vaccinated and 1:1 propensity-matched unvaccinated cohorts, and corresponding vaccine efficacy. Incidence is calculated as the number of cases per 1000 person-days. The columns are: **(1) Time Period:** Time period relative to first vaccine dose for vaccinated cohort or study enrollment day for unvaccinated cohort; **(2) Vaccinated Incidence Rate:** Number of patients with positive PCR tests in the vaccinated cohort in the time period, divided by the number of at-risk person-days for the vaccinated cohort in the time period; in brackets, the number of cases per 1000 person-days; **(3) Unvaccinated Incidence Rate:** Number of patients with positive PCR tests in the propensity-matched unvaccinated cohort in the time period, divided by the number of at-risk person-days for the propensity-matched unvaccinated cohort in the time period; in brackets, the number of cases per 1000 person-days; **(4) Incidence Rate Ratio:** Vaccinated Incidence Rate divided by Unvaccinated Incidence Rate, along with the exact 95% confidence interval (17), **(5) Vaccine Efficacy:** 100% x (1-Incidence Rate Ratio), along with the 95% confidence interval.

| **Time Period** | **Vaccinated Incidence Rate**<br>Cases/Person-Days<br>[Per 1000 Person-Days]<br>(# Patients Contributing) | **Unvaccinated Incidence Rate**<br>Cases/Person-Days<br>[Per 1000 Person-Days]<br>(# Patients Contributing) | **Incidence Rate Ratio**<br>(95% CI) | **Vaccine Efficacy**<br>(95% CI) |
| --- | --- | --- | --- | --- |
| Day 1 onwards | 263 / 835,535<br>[0.31]<br>(n = 31,069) | 661 / 825,106<br>[0.8]<br>(n = 31,069) | 0.39 (0.34, 0.45) | 60.7%<br>(54.6%, 66.1%) |
| Day 15 onwards | 69 / 451,783<br>[0.15]<br>(n = 21,712) | 271 / 443,207<br>[0.61]<br>(n = 21,497) | 0.25 (0.19, 0.33) | 75.0%<br>(67.4%, 81.1%) |
| Day 29 onwards | 14 / 175,549<br>[0.08]<br>(n = 15,482) | 81 / 170,781<br>[0.47]<br>(n = 15,179) | 0.17 (0.088, 0.3) | 83.2% (70.1%, 91.2%) |
| Day 36 onwards | 6 / 84,638<br>[0.071]<br>(n = 8,263) | 35 / 81,873<br>[0.43]<br>(n = 8,046) | 0.17 (0.057, 0.4) | 83.4%<br>(60.2%, 94.3%) |
| Day 36 onwards<br>(2 doses) | 4 / 82,686<br>[0.048]<br>(n = 8,041) | 35 / 81,873<br>[0.43]<br>(n = 8,046) | 0.11 (0.029, 0.32) | 88.7%<br>(68.4%, 97.1%) |
| Days 1-7 | 100 / 208,496<br>[0.48]<br>(n = 31,069) | 215 / 207,877<br>[1.0]<br>(n = 31,069) | 0.46 (0.36, 0.59) | 53.6%<br>(40.9%, 63.8%) |
| Days 8-14 | 94 / 175,256<br>[0.54]<br>(n = 26,750) | 175 / 174,022<br>[1.0]<br>(n = 26,611) | 0.53 (0.41, 0.69) | 46.7%<br>(31.1%, 58.9%) |
| Days 15-21 | 33 / 147,115<br>[0.22]<br>(n = 21,712) | 106 / 145,382<br>[0.73]<br>(n = 21,497) | 0.31 (0.2, 0.46) | 69.2%<br>(54.1%, 79.8%) |
| Days 22-28 | 22 / 129,119<br>[0.17]<br>(n = 19,634) | 84 / 127,044<br>[0.66]<br>(n = 19,361) | 0.26 (0.15, 0.42) | 74.2%<br>(58.4%, 84.7%) |
| Days 29-35 | 8 / 90,911<br>[0.088]<br>(n = 15,482) | 46 / 88,908<br>[0.52]<br>(n = 15,179) | 0.17 (0.069, 0.36) | 83.0%<br>(63.6%, 93.1%) |
| Days 36-42 | 2 / 53,300<br>[0.038]<br>(n = 8,263) | 26 / 51,773<br>[0.5]<br>(n = 8,046) | 0.075 (0.0086, 0.3) | 92.5%<br>(70.2%, 99.1%) |

**Figure 2.**
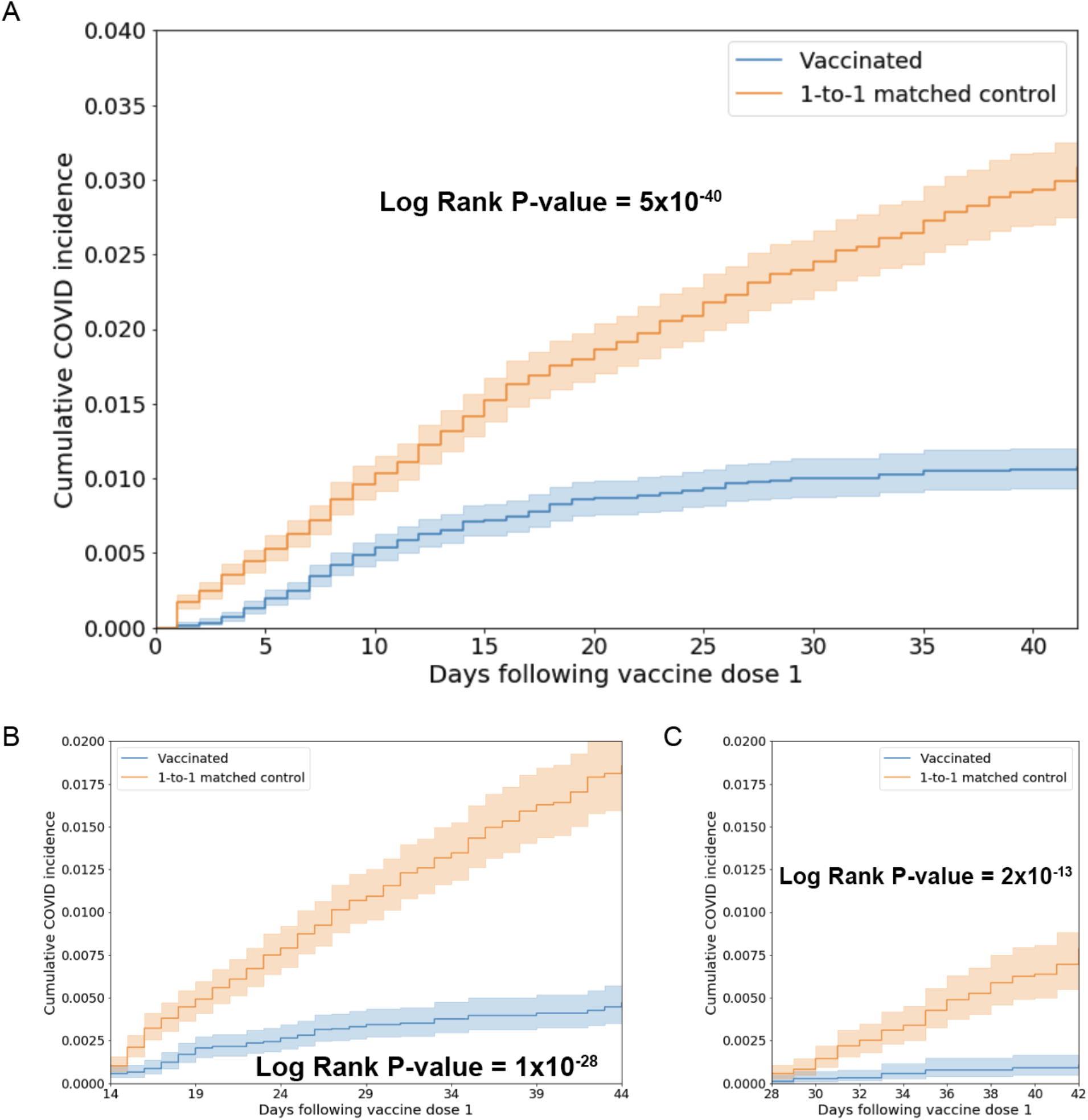
Kaplan Meier analyses to assess cumulative proportional incidence of SARS-CoV-2 infection between vaccinated and unvaccinated individuals. Cumulative proportional incidence at time *t* is the estimated proportion of patients who experience the outcome on or before time *t*, i.e. 1 minus the standard Kaplan-Meier survival estimate. Cumulative proportional incidence of SARS-CoV-2 infection with onset on any day after the date of first vaccination (A), after 14 days from the date of first vaccination (B), or after 28 days from the date of first vaccination (C), against a 1-to-1 matched unvaccinated control cohort. A log-rank test rejects the null hypothesis of equal hazard rates with p-values of 5×10^−40^ (A), 1×10^−28^ (B), and 2.4×10^−13^ (C). Note that the x-axis of (B) ranges from 14 to 42 days (following the first vaccine dose), and the x-axis of (C) ranges from 28 to 42 days (following the first vaccine dose).

**Figure 3.**
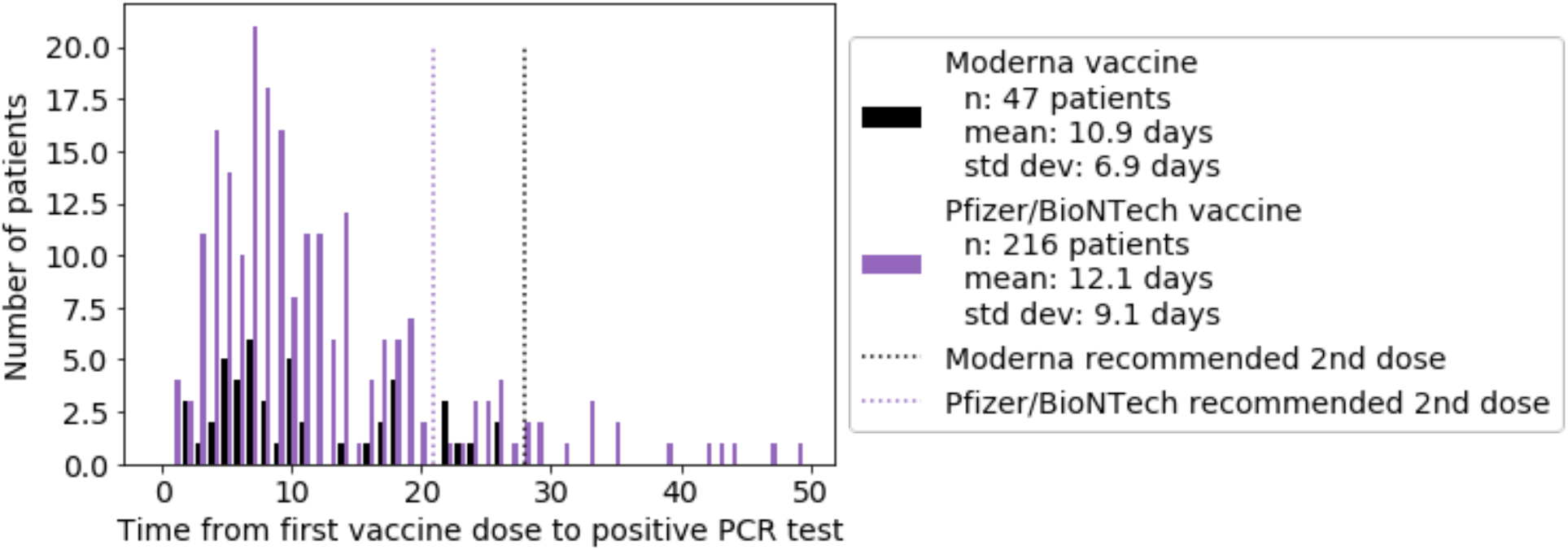
Distribution of the time from first vaccine dose to first positive PCR test, for the patients with at least one positive PCR test following vaccination. Patient counts for mRNA-1273 (Moderna vaccine) are shown in black, and patient counts for BNT162b2 (Pfizer/BioNTech vaccine) are shown in purple. For mRNA-1273, the mean time to positive PCR test following the first dose is 10.9 days (standard deviation: 6.9 days), and for BNT162b2, the mean time to positive PCR test following the first dose is 12.1 days (standard deviation: 9.1 days). Dotted lines indicate the recommended time for the second vaccine dose for mRNA-1273 (28 days) and BNT162b2 (21 days).

Starting 36 days after study enrollment (approximately two weeks after the second dose of BNT162b2 and one week after the second dose of mRNA-1273), the vaccinated and unvaccinated incidence rates were 0.071 and 0.43 case per 1000 person-days, respectively. This corresponds to a vaccine efficacy of 83.4% (95% CI: 60.2-94.3%) (**Table 2**). Importantly, we found that two of the six infections in the vaccinated cohort on or after day 36 occurred in individuals who had received only one vaccine dose, even though all vaccinated individuals should have received two doses by this time point per the manufacturer guidelines. Among the properly vaccinated individuals (i.e. those who had received both doses prior to day 36), the incidence rate of a positive SARS-CoV-2 PCR test was 0.048 cases per 1000 person-days (4 cases in 82,686 person days), indicating an efficacy of 88.7% (95% CI: 68.4-97.1%) (**Table 2**).

We also assessed the rates of infection and estimated vaccine efficacy in six non-overlapping 7-day intervals starting at the date of first vaccination. Even in the first seven days after study enrollment, vaccinated individuals had significantly lower infection incidence rates (0.48 cases per 1000 person-days) than unvaccinated individuals (1.0 cases per 1000 person-days), corresponding to an efficacy of 53.6% (95% CI: 40.9%-63.8%) (**Table 2**). The vaccination efficacy then generally increased in subsequent weeks, reaching its maximum (92.5%; 95% CI: 70.2-99.1%) during the sixth week after study enrollment (days 36-42) (**Table 2**).

### COVID-19 vaccines are associated with lower hospitalization rates post SARS-CoV-2 infection

To assess whether vaccination also reduces illness severity, we compared 14-day rates of hospitalization, ICU admission, and mortality in COVID-19 patients who were vaccinated prior to diagnosis (n = 263) and 1-to-10 propensity score matched unvaccinated COVID-19 patients (n = 2,630) (see **Methods** and **Table 3**). The vaccinated population showed a significantly lower 14-day hospital admission rate (3.7% vs. 9.2%; Relative Risk = 0.4; p = 0.0074) (**Figure 4A, Table 4**). On the other hand, ICU admission rates were similar between these cohorts (1% vs. 1.3%; Relative Risk = 0.82; p = 1.0) (**Figure 4B, Table 4**). 14-day conditional mortality rates were also not significantly different (0% vs. 0.085%; Relative Risk = 0; p = 1.0), but it is worth noting that no vaccinated patients died within 14 days of acquiring COVID-19 (**Table 4**). In fact, none of the vaccinated patients who were subsequently diagnosed with COVID-19 have died, including 59 with at least 28 days of follow-up (data not shown).

**Table 4.** 14-day rates of hospitalization, ICU admission, and mortality for vaccinated vs 1:10 propensity-matched unvaccinated COVID-19 patients. Patients were considered eligible for analysis if they had at least 14 days of follow-up after COVID-19 diagnosis as defined by a positive SARS-CoV-2 PCR test. For each outcome, the relative risk (and its 95% confidence interval) and Fisher exact test p-value are used to compare the rates between vaccinated and unvaccinated patients. To indicate statistical significance, * denotes p-value < 0.05, ** denotes p-value < 0.01.

| **Outcome** | **Moderna or Pfizer 1+ dose, COVID positive (263 patients)** | **Matched unvaccinated, COVID positive (2630 patients)** | **Relative Risk (95% CI)** | **Fisher Exact test p-value** |
| --- | --- | --- | --- | --- |
| Number of patients with at least 14 days of follow-up | 191 | 2348 |  |  |
| 14-Day Hospital admission rate | 7 / 191 (3.7%) | 217 / 2348 (9.2%) | 0.4 (0.21, 0.86) | 0.0074** |
| 14-Day ICU admission rate | 2 / 191 (1%) | 30 / 2348 (1.3%) | 0.82 (0.28, 3.6) | 1 |
| 14-Day Mortality rate | 0 / 191 (0%) | 2 / 2348 (0.085%) | 0 (0, 51) | 1 |

**Figure 4.**
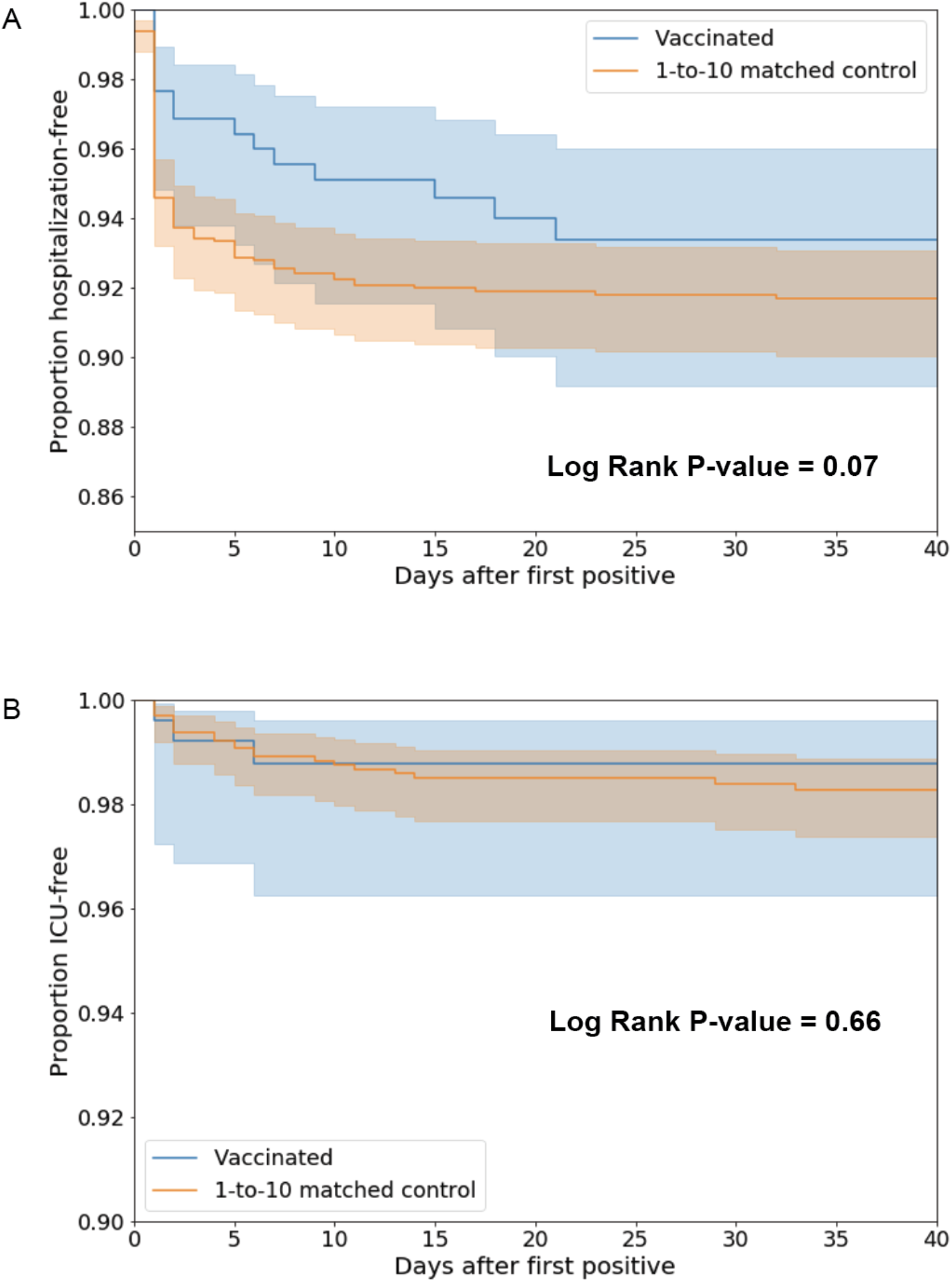
Kaplan Meier analyses to assess COVID-19 disease severity between vaccinated and unvaccinated patients. (A) Hospitalization-free survival comparison between patients who tested positive for SARS-CoV-2 after being vaccinated versus 1:10 propensity-matched patients who tested COVID-positive and were not vaccinated. A log-rank test fails to reject the null hypothesis of equal hazard rates with a p-value of 0.07. (B) ICU-free survival comparison between patients who tested positive for SARS-CoV-2 after being vaccinated versus 1:10 propensity-matched patients who tested COVID-positive and were not vaccinated. A log-rank test fails to reject the null hypothesis of equal hazard rates with a p-value of 0.66.

## Discussion

Recent phase 3 trials have led to the authorization of two COVID-19 vaccines in the United States, and other vaccines have been approved in other countries or show promise for approval in the near future (7, 8). This study provides strong further evidence supporting the use of vaccination to prevent and reduce the severity of COVID-19. While other real world analyses of COVID-19 vaccines are now emerging (22), a defined placebo group or adequately balanced unvaccinated cohort is difficult to ascertain outside of the clinical trial setting. To address this challenge, we used propensity matching to generate cohorts of vaccinated and unvaccinated patients who are balanced for demographic, geographic, and social variables, and then evaluated the effect of vaccination on the rate of SARS-CoV-2 positivity and COVID-19 severity between these cohorts. These vaccines, when administered as two serial doses, were 88.7% effective (95% CI: 68.4-97.1%) in preventing SARS-CoV-2 infection with onset at least 36 days after the first dose. This result is in line with the previously reported efficacies for both BNT162b2 (95.0%; 95% CI: 90.3-97.6%) and mRNA-1273 (94.1%; 95% CI: 89.3-96.8%) in preventing symptomatic COVID-19 with onset at least 28 or 42 days after the first dose, respectively (9, 10).

That the efficacy observed in our study is slightly lower than those reported in the two corresponding randomized controlled trials should be interpreted cautiously and contextually, as there are several plausible reasons for this. First, the 95% confidence intervals of all three efficacy estimates are highly overlapping, consistent with the true efficacies not being meaningfully different from each other. Second, due to distribution guidelines that are in place for Phase 1a of the vaccine rollout (11), individuals at high risk for acquiring COVID-19 (e.g., healthcare workers and residents of long term care facilities) are expected to be overrepresented in this vaccinated cohort. This could lead to an underestimation of vaccine efficacy, as the propensity matched unvaccinated group is likely composed of lower-risk individuals despite being matched for age, sex, race, ethnicity, and the number of prior SARS-CoV-2 PCR tests. Third, the likelihood of exposure to SARS-CoV-2 may be dependent on vaccination status to a greater extent in the real world than it is in the context of a randomized trial. For example, vaccinated individuals may feel more comfortable participating in social situations that pose a higher risk for infection, whereas this bias did not exist by definition in the context of the observer-blinded clinical trials. On the other hand, vaccinated individuals may be more inclined to take precautionary measures such as wearing masks and washing hands frequently in order to reduce their risk of infection, which could result in the “healthy user bias” (23). Follow-up observational studies are required to assess the associations between COVID-19 vaccination and health-seeking behaviors such as adherence to COVID-19 social distancing guidelines.

The incidence rates of SARS-CoV-2 infection in both our vaccinated and unvaccinated cohorts (113 and 292 cases per 1000 person-years, respectively) are notably higher than the COVID-19 incidence rates reported in the placebo groups of both the BNT162b2 and mRNA-1273 trials (69.1 and 79.7 cases per 1000 person-years, respectively), but there are also several explanations for this observation. First and most importantly, in contrast to both clinical trials, our outcome of interest is only a positive SARS-CoV-2 PCR test (with no requirement for presence of any clinical symptoms), whereas the phase 3 trials were designed to study symptomatic COVID-19 infections. Given that over 40% of COVID-19 cases may be asymptomatic (24, 25), we know that the rates of SARS-CoV-2 PCR positivity would have been higher than the rates of symptomatic COVID-19 in both trials. This discrepancy in measured outcomes may also contribute to the slight differences in estimated efficacy addressed previously. Second, due to overrepresentation of high-risk individuals in the vaccinated cohort as described above, it is reasonable to hypothesize that the infection incidence was positively skewed in this study.

Another interesting result from this study is the trend in vaccine efficacy over time. We observe that the estimated efficacy for Days 1-7 is 53.6% (95% CI: [40.9%, 63.8%]), while the estimated efficacy for Days 8-14 is 46.7% (95% CI: [31.1%, 58.9%]). One explanation could be that some individuals who initially presented with symptoms for COVID-19 were filtered out from the vaccinated cohort. This could explain the low rates of SARS-CoV-2 infection observed in the first week following the first vaccine dose. As a result, we should use caution when interpreting the results from the time periods “Days 1-7” and “Days 1 onwards” in this study.

Our finding that hospitalization rates are lower in COVID-19 patients who were vaccinated prior to SARS-CoV-2 infection compared to propensity matched unvaccinated COVID-19 patients is consistent with the clinical trial results for both BNT162b2 and mRNA-1273 (9, 10). In the trial of BNT162b2, 9 cases of severe COVID-19 occurred in the placebo group compared to only one in the vaccinated cohort; and in the trial of mRNA-1273, 30 cases of severe COVID-19 occurred, all of which were in the placebo group. While the ICU admission and mortality rates were not significantly lower in our vaccinated population, this may be attributable to an inadequate number of patients with these outcomes in either group to date in our study. As more patients are vaccinated and follow-up time increases, we will update our analyses to determine whether vaccination can also reduce the risk of these outcomes.

There are several important limitations to consider in this study. First, while the cohort size was even larger than the cohorts studied in phase 3 trials, the mean follow-up time per patient is substantially lower (mean = 27.1 days versus approximately 80 to 90 days). Consistent with this, approximately 45.1% of our vaccinated cohort had received only one dose of vaccination at the time of this study (**Table S1**). We were thus limited in the number of patients and at-risk person-days that were available for the critical long term efficacy analyses. Second, we did not assess vaccine-associated adverse events, nor did we compare the clinical symptomatology of COVID-19 infections between vaccinated and unvaccinated patients. Third, it is possible that the likelihood of seeking out a SARS-CoV-2 PCR test was different between vaccinated and propensity matched unvaccinated patients, which could introduce bias into our estimates of vaccine efficacy. Indeed, vaccinated patients may feel less compelled to undergo subsequent PCR testing, thereby reducing the number of positive tests recorded in this group. However, our data suggests that this is likely not a strong confounding factor, as the fraction of vaccinated patients with at least one PCR test after study enrollment (13.9%) was only marginally lower than the same fraction of unvaccinated patients (16.6%) (**Table S1**), and the cumulative counts of PCR tests are similar over time for the vaccinated and unvaccinated cohorts (**Figure S3**). Finally, the cohorts utilized to assess vaccine efficacy were predominantly female (vaccinated: 62.2%; unvaccinated: 64.8%). As vaccine distribution expands in the coming months, we will be able to analyze cohorts which are more balanced for sex and to robustly assess vaccine efficacy in males and females separately.

Our data demonstrates a strong real world effect of COVID-19 vaccination on par with the results reported in each randomized trial. This study also provides additional information which could not be ascertained from either trial, including the conclusions that (1) vaccination is effective in individuals who are at highest risk for acquiring COVID-19, and (2) vaccination reduces the rate of SARS-CoV-2 infection as defined by a positive PCR test alone. In summary, we emphasize that COVID-19 vaccines should be administered as broadly and rapidly as possible to the public and that the real world efficacy of these vaccines should be continuously monitored as we move beyond Phase 1a of the distribution process.

## Data Availability

After publication, the data will be made available to others upon reasonable requests to the corresponding author. A proposal with a detailed description of study objectives and the statistical analysis plan will be needed for evaluation of the reasonability of requests. Deidentified data will be provided after approval from the corresponding author and the Mayo Clinic.

## Supplementary Material

**Figure S1.**
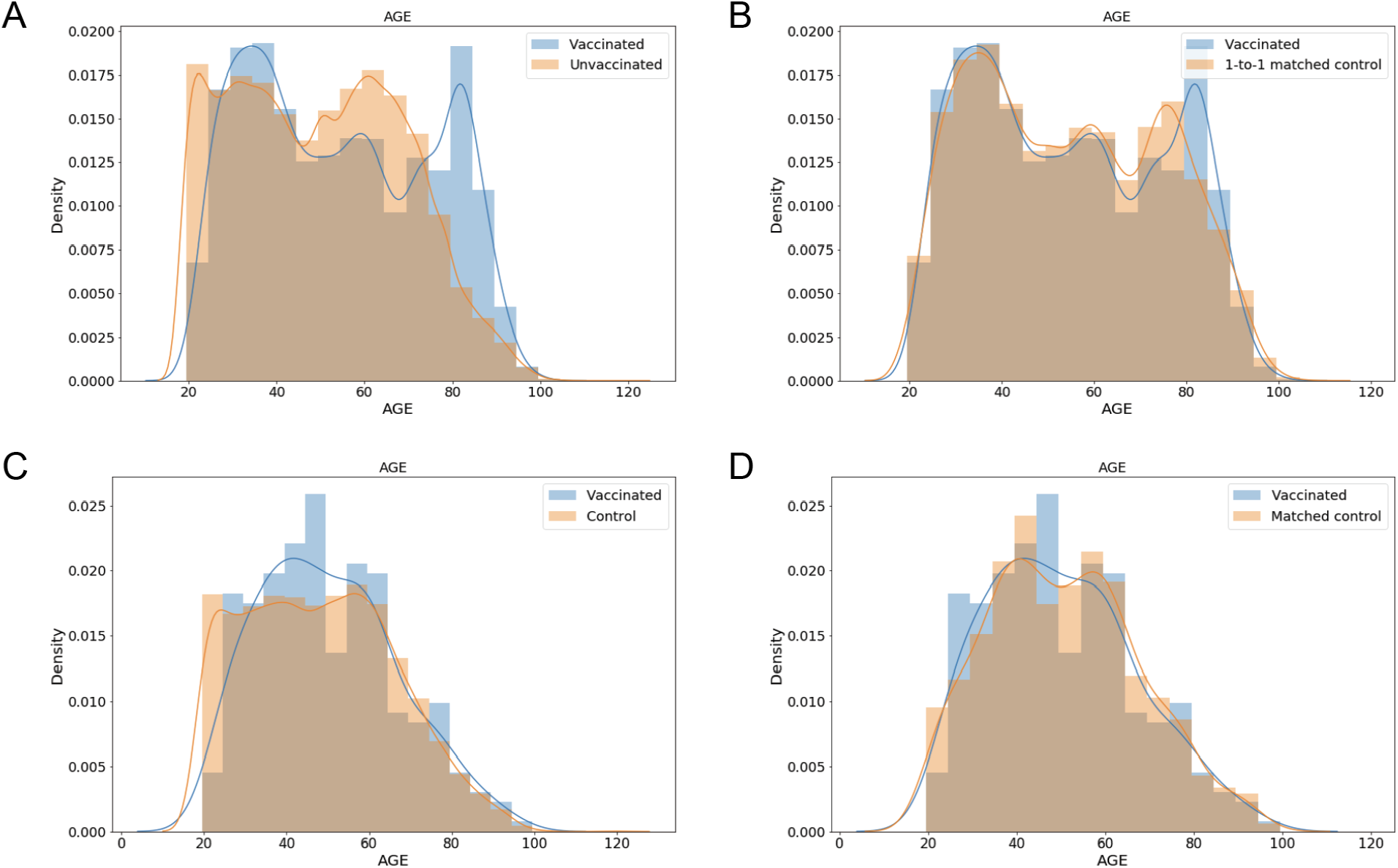
Age distributions for vaccinated and unvaccinated cohorts before and after propensity matching. (A-B) Distribution of ages for vaccinated patients and unvaccinated patients before (A) and after (B) 1-to-1 matching. These matched cohorts were used to assess vaccine efficacy in preventing SARS-CoV-2 infection. (C-D) Distribution of ages for COVID-19 patients who were vaccinated prior to diagnosis and unvaccinated COVID-19 patients before (C) and after (D) 1-to-10 propensity score matching. These matched cohorts were used to assess the effect of vaccination on COVID-19 disease severity.

**Figure S2.**
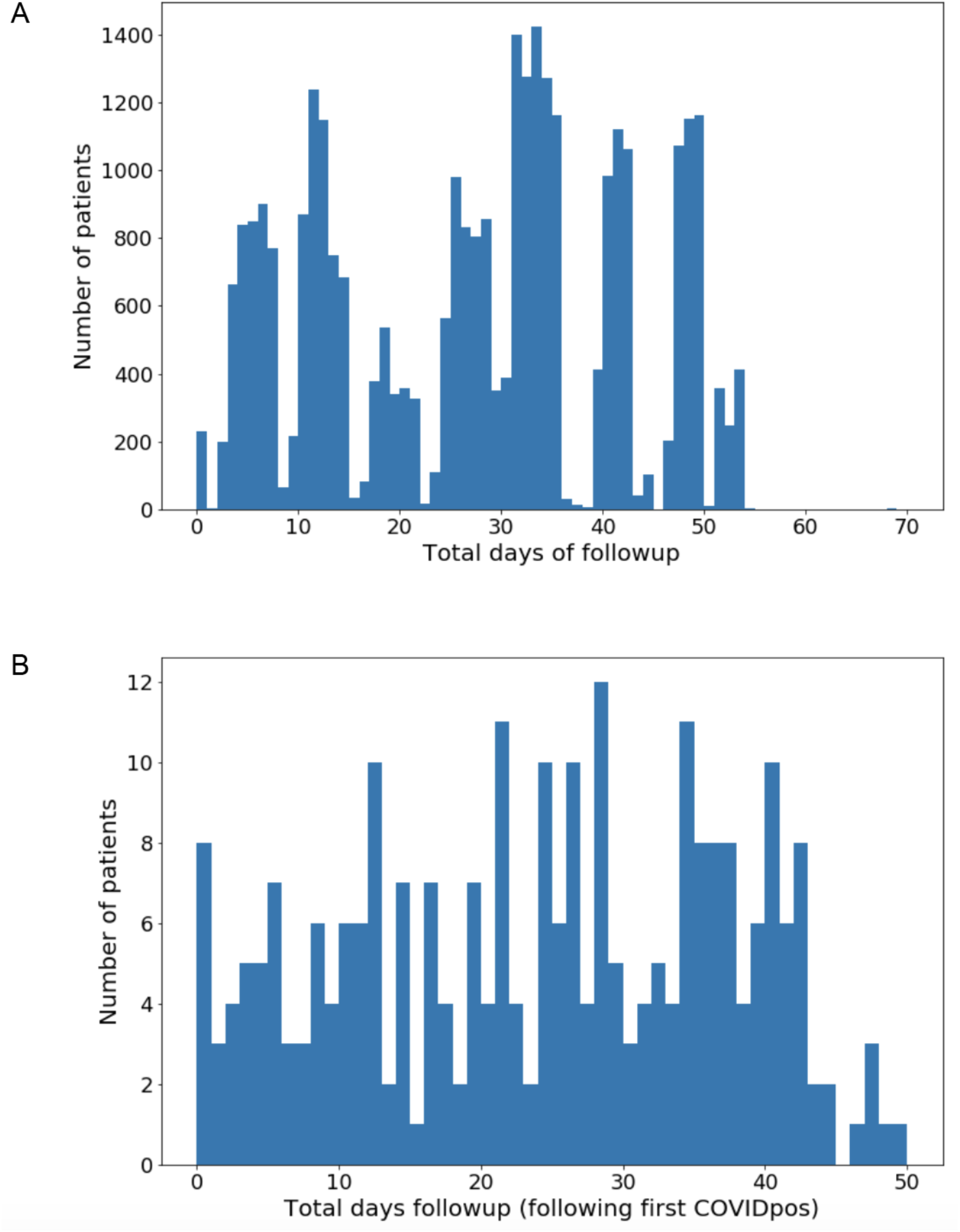
Distributions of total available follow-up time for study cohorts of interest. (A) Total available follow-up time (days) for the 31,299 individuals who received a COVID-19 vaccine and did not have a positive SARS-CoV-2 test before their first vaccine dose. Total available follow-up time is defined as the number of days from first vaccine to the final study date (February 8, 2020), including days after COVID-19 diagnosis or death, if applicable. (B) Total available follow-up time (days) for the 263 individuals who received a COVID-19 vaccine subsequently tested positive for SARS-CoV-2 by PCR. Total available follow-up time is defined as the number of days from diagnosis (date of positive SARS-CoV-2 test) to the final study date (February 8, 2020), including days after COVID-19 diagnosis or death, if applicable.

**Figure S3.**
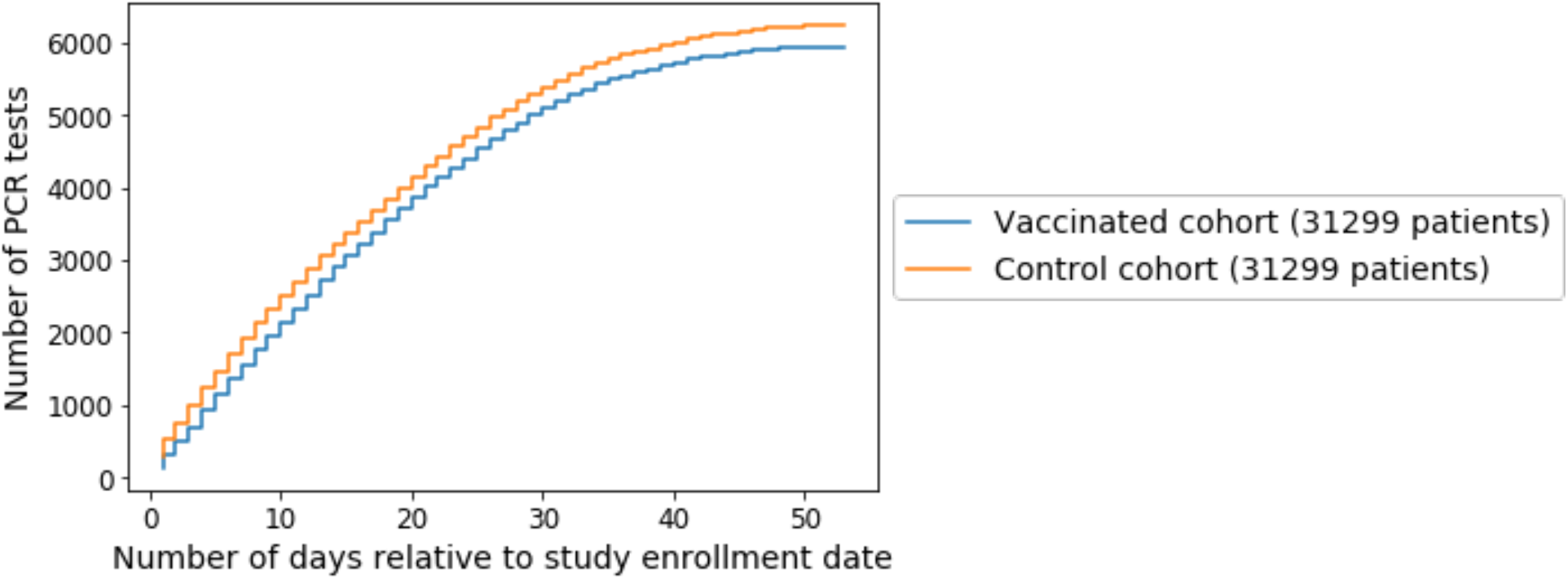
Cumulative number of PCR tests for the propensity matched vaccinated and control cohorts. The x-axis corresponds to the number of days relative to the study enrollment date. For the vaccinated cohort, the study enrollment date is the date of the first vaccine dose. For the control cohort, the study enrollment date is the date of the first vaccine dose for the matched vaccinated individual. The y-axis corresponds to the total number of PCR tests taken by the vaccinated (**blue**) or control (**orange**) cohorts.

**Table S1.**
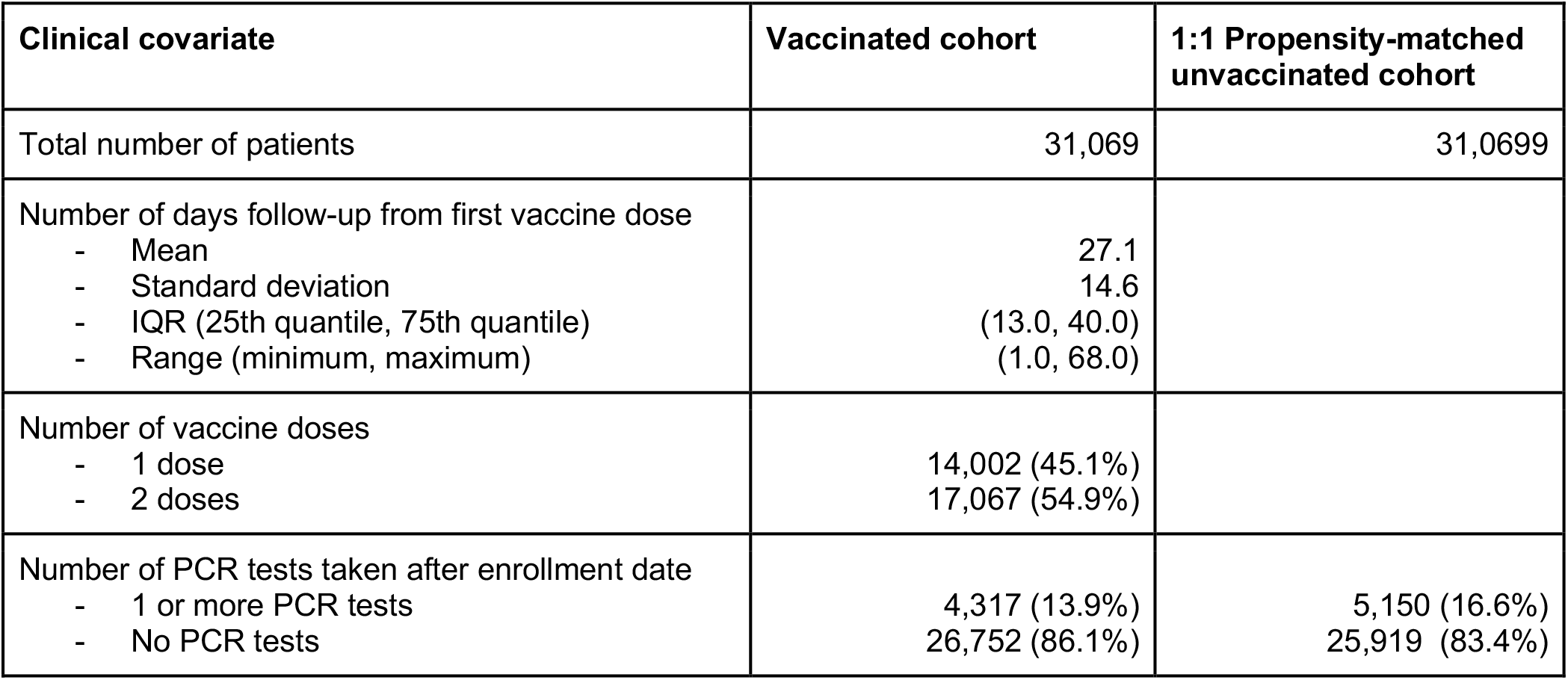
Follow-up, dosing, and PCR testing information for the vaccinated and 1:1 propensity-matched unvaccinated cohorts. For the vaccinated cohort, descriptive statistics on the number of doses received and the number of days of follow-up from the first vaccine dose are provided. For both cohorts, the number of PCR tests after the enrollment date are provided. For the vaccinated cohort, the enrollment date corresponds to the date of the first vaccine dose. For the 1:1 propensity-matched cohort, the enrollment date corresponds to the date of the first vaccine dose for the matched vaccinated patient.

| Clinical covariate | Vaccinated cohort | 1:1 Propensity-matched unvaccinated cohort |
| --- | --- | --- |
| Total number of patients | 31,069 | 31,0699 |
| Number of days follow-up from first vaccine dose<br>- Mean<br>- Standard deviation<br>- IQR (25th quantile, 75th quantile)<br>- Range (minimum, maximum) | 27.1<br>14.6<br>(13.0, 40.0)<br>(1.0, 68.0) |  |
| Number of vaccine doses<br>- 1 dose<br>- 2 doses | 14,002 (45.1%)<br>17,067 (54.9%) |  |
| Number of PCR tests taken after enrollment date<br>- 1 or more PCR tests<br>- No PCR tests | 4,317 (13.9%)<br>26,752 (86.1%) | 5,150 (16.6%)<br>25,919 (83.4%) |

## References

1. COVID-19 Map - Johns Hopkins Coronavirus Resource Center. at <https://coronavirus.jhu.edu/map.html>

2. Lan, J. et al. Structure of the SARS-CoV-2 spike receptor-binding domain bound to the ACE2 receptor. Nature 581, 215–220 (2020).

3. Shang, J. et al. Structural basis of receptor recognition by SARS-CoV-2. Nature 581, 221– 224 (2020).

4. Venkatakrishnan, A. J. et al. Knowledge synthesis of 100 million biomedical documents augments the deep expression profiling of coronavirus receptors. (2020). doi:10.7554/eLife.58040

5. Jackson, L. A. et al. An mRNA Vaccine against SARS-CoV-2 - Preliminary Report. N. Engl. J. Med. 383, (2020).

6. Mulligan, M. J. et al. Phase I/II study of COVID-19 RNA vaccine BNT162b1 in adults. Nature 586, 589–593 (2020).

7. Voysey, M. et al. Safety and efficacy of the ChAdOx1 nCoV-19 vaccine (AZD1222) against SARS-CoV-2: an interim analysis of four randomised controlled trials in Brazil, South Africa, and the UK. Lancet 397, (2021).

8. Sadoff, J. et al. Interim Results of a Phase 1-2a Trial of Ad26.COV2.S Covid-19 Vaccine. N. Engl. J. Med. (2021). doi:10.1056/NEJMoa2034201

9. Polack, F. P. et al. Safety and Efficacy of the BNT162b2 mRNA Covid-19 Vaccine. N. Engl. J. Med. 383, 2603–2615 (2020).

10. Baden, L. R. et al. Efficacy and Safety of the mRNA-1273 SARS-CoV-2 Vaccine. N. Engl. J. Med. (2020). doi:10.1056/NEJMoa2035389

11. CDC. CDC’s COVID-19 Vaccine Rollout Recommendations. (2021). at <https://www.cdc.gov/coronavirus/2019-ncov/vaccines/recommendations.html>

12. Austin, P. C. An Introduction to Propensity Score Methods for Reducing the Effects of Confounding in Observational Studies. Multivariate Behav. Res. 46, 399–424 (2011).

13. Austin, P. C. A comparison of 12 algorithms for matching on the propensity score. Stat. Med. 33, 1057–1069 (2014).

14. Austin, P. C. Balance diagnostics for comparing the distribution of baseline covariates between treatment groups in propensity-score matched samples. Stat. Med. 28, 3083 (2009).

15. Stuart, E. A., Lee, B. K. & Leacy, F. P. Prognostic score–based balance measures for propensity score methods in comparative effectiveness research. J. Clin. Epidemiol. 66, S84 (2013).

16. J Martin Bland, D.G.A. Statistics Notes: The logrank test. BMJ : British Medical Journal 328, 1073 (2004).

17. Sahai, H. & Khurshid, A. Statistics in Epidemiology: Methods, Techniques and Applications. (CRC Press, 1995).

18. CDC. COVID-19 and Your Health. (2020). at<https://www.cdc.gov/coronavirus/2019-ncov/need-extra-precautions/evidence-table.html>

19. Miguel Angel Luque Fernandez; Michael Schomaker; Sho Komukai; Aurelien Belot; Daniel Redondo-Sanchez; Bernard Rachet; Mireille Schnitzer & Miguel Angel Luque Fernandez, MA, MPH, MSc. Delta Method in Epidemiology: An Applied and Reproducible Tutorial. at <https://migariane.github.io/DeltaMethodEpiTutorial.nb.html>

20. Devlin, J., Chang, M.-W., Lee, K. & Toutanova, K. BERT: Pre-training of Deep Bidirectional Transformers for Language Understanding. (2018). at <http://arxiv.org/abs/1810.04805>

21. Wagner, T. et al. Augmented curation of clinical notes from a massive EHR system reveals symptoms of impending COVID-19 diagnosis. Elife 9, (2020).

22. Rossman, H. et al. Patterns of COVID-19 pandemic dynamics following deployment of a broad national immunization program. medRxiv 2021.02.08.21251325 (2021).

23. Shrank, W. H., Patrick, A. R. & Alan Brookhart, M. Healthy User and Related Biases in Observational Studies of Preventive Interventions: A Primer for Physicians. J. Gen. Intern. Med. 26, 546 (2011).

24. Oran, D. P. & Topol, E. J. Prevalence of Asymptomatic SARS-CoV-2 Infection : A Narrative Review. Ann. Intern. Med. 173, (2020).

25. Sayampanathan, A. A. et al. Infectivity of asymptomatic versus symptomatic COVID-19. Lancet 397, (2021).

